# Explainable advanced electrocardiography at rest for ruling out myocardial ischemia on stress echocardiography

**DOI:** 10.1101/2025.01.24.25320506

**Authors:** Kevin X Yang, Johan von Scheele, Maren Maanja, Daniel E Loewenstein, Todd T Schlegel, Martin Ugander, Rebecca Kozor

## Abstract

**Background:** Stress echocardiography (SE) is relatively resource intensive and has a low incidence of abnormal tests for detecting coronary artery disease (CAD) in low-to-intermediate risk patients. This study aimed to derive and determine the diagnostic performance of a resting advanced electrocardiography (A-ECG) score for detecting inducible myocardial ischemia on SE in patients with low-to-intermediate risk stable chest pain.

**Methods:** Patients were included if they presented with low-to-intermediate risk stable chest pain to the emergency department, had acute coronary syndrome ruled out by electrocardiography (ECG) and high-sensitivity troponin, and subsequently underwent outpatient SE. Patients were excluded if they had known CAD or confounders on resting ECG. A-ECG was retrospectively applied to a standard resting 12-lead ECG and a multivariable logistic regression score was derived to predict myocardial ischemia on SE.

**Results:** Among 292 patients (51% male, age 58±14 years), 24 (8%) exhibited inducible myocardial ischemia on SE. A 3-parameter A-ECG score had an area under the receiver-operating characteristic curve (AUC [bootstrapped 95% confidence interval]) of 0.85 [0.75– 0.93], sensitivity 92 [67–100]%, specificity 67 [64–94]%, positive predictive value 22 [20**–**55]%, negative predictive value 99 [96**–**100]%, positive likelihood ratio 2.8 [2.5–12.0] and inverse negative likelihood ratio 8.1 [2.5-18.0].

**Conclusions:** An A-ECG score had a good overall diagnostic performance and excellent performance for ruling out inducible myocardial ischemia on SE. This supports the use of an A-ECG score to triage and improve the selection of patients with low-intermediate risk stable chest pain that should undergo further testing with SE.

## Introduction

Stress echocardiography (SE) is well-validated for diagnostic and prognostic use in coronary artery disease (CAD)^1,2^. It is routinely performed to investigate stable chest pain in patients with a low-intermediate risk of CAD. However, SE is relatively time and resource-consuming, requiring considerable expertise for reliable interpretation. The incidence of inducible myocardial ischemia on SE and other stress imaging modalities has also declined over time, now as low as 4% in contemporary populations^3,4^. This incurs unnecessary costs to patients and the healthcare system without substantial clinical utility. As such, there is an unmet need for a triage tool that more accurately predicts which patients will exhibit inducible myocardial ischemia.

The standard resting 12-lead electrocardiogram (ECG) is simple, low-cost, quickly performed, and widely available. It is ubiquitously conducted for patients presenting to the emergency department with chest pain. However, visual assessment of the resting ECG has poor diagnostic accuracy for detecting CAD^5^. Advanced electrocardiography (A-ECG) involves the measurement of various advanced and conventional ECG measures and subsequently using multivariable statistics to detect heart disease^6^. A-ECG incorporates signal averaging, derived vectorcardiography, and measurement of QRS and T waveform complexity by singular value decomposition^7,8^. A-ECG analysis can be applied to a standard resting 12-lead digital ECG, providing diagnostic information that cannot be discerned by visual assessment. In one study, A-ECG was shown to rule out significant CAD based on cardiac computed tomography angiography (CCTA) with high specificity^9^. This supports its use as a powerful triaging tool for chest pain by identifying patients who do not require further investigations. However, A-ECG has not yet been validated using SE.

Therefore, the aim of the study was to derive and determine the diagnostic performance of an A-ECG score for detecting inducible myocardial ischemia on SE in patients with low-to-intermediate risk stable chest pain. We hypothesised that an A-ECG score from the resting 12-lead ECG would have a clinically useful diagnostic performance for detecting inducible myocardial ischemia on SE in these patients.

## Methodology

### Study participants

The study population consisted of adult patients retrospectively recruited between February 2017 and March 2022 who presented with cardiac-suspected chest pain to the emergency department at Royal North Shore Hospital, had myocardial infarction ruled out by resting ECG and blood troponin levels, were deemed low-to-intermediate risk by HEART score^10^, and were subsequently referred to the rapid access chest pain clinic (RACC) for SE. Patients were excluded if they had a pacemaker or known CAD defined as having a history of myocardial infarction, percutaneous coronary intervention, coronary artery bypass grafting or documented coronary artery stenosis >50% on invasive angiography or CCTA. Patient data was retrospectively obtained from electronic medical records. The study was approved by the Northern Sydney Local Health District Human Research Ethics Committee (2019/STE10454). All patients provided written informed consent, or a waiver of consent was provided by the ethics committee.

### Stress echocardiography

All SE tests were performed using an exercise treadmill following the Bruce protocol^11^. Inducible myocardial ischemia on SE was defined as a stress-induced left ventricular regional wall motion abnormality as determined by the reporting cardiologist in the RACC. Patients were excluded if they had an uninterpretable test or if their peak heart rate was less than 85% of their maximum predicted heart rate (220 – age) without ischemia.

### Exercise electrocardiography

Treadmill exercise ECG (ExECG) was performed simultaneously alongside SE in all patients. Inducible myocardial ischemia on ExECG was defined as ≥ 1 millimetre of horizontal or down-sloping ST depression in 2 contiguous leads during exercise or recovery. All ST segment measurements were performed 80 milliseconds after the J-point. Patients who tested negative on SE but had equivocal findings on ExECG (horizontal/down-sloping ST segment depression < 1 millimetre) were excluded.

### Resting ECG Acquisition and Analysis

A 10-second resting digital ECG was performed (Philips, Best, the Netherlands) during RACC visit prior to SE. Patients were excluded if their digital ECG could not be retrospectively retrieved. A large proportion of digital ECGs were not stored due to technical issues in the early establishment of the RACC. Available ECG files were anonymized, reviewed, and excluded if they contained atrial fibrillation or atrial flutter, QRS duration > 120 milliseconds, missing leads, or excessive noise. Additionally, a large proportion of files contained 50 Hz noise from electrical interference which was not detectable at the time of recording due to a noise filter, and needed to be excluded. Included ECGs were analysed using in-house software to produce conventional and advanced ECG parameters. Conventional parameters included all major intervals, axes, voltages and ST segment levels. Advanced parameters included derived vectorcardiographic measures and waveform complexity measures of the QRS and T wave quantified by singular value decomposition.

### Statistical Analysis

All statistical analyses were performed using the programming language R 4.1.2 (R Core Team, R Foundation for Statistical Computing, Vienna, Austria). Categorical variables are expressed as frequencies and percentages. Continuous variables are expressed as mean±standard deviation. Baseline characteristics between groups were compared using Fisher’s exact test for categorical variables and one-way analysis of variance for continuous variables. A multivariable logistic regression model was trained for inducible myocardial ischemia on SE. Feature selection was performed by first identifying univariately significant parameters using the Mann-Whitney U test. Exhaustive search was then performed on these parameters to identify the best performing model, with the number of parameters included in the model limited to 3 (∼1 parameter per 10 events). Performance was measured using the area under the receiver operating characteristic curve (AUC), sensitivity, specificity, predictive values, and likelihood ratios. The multivariate statistical significance of model parameters were evaluated using the likelihood-ratio chi-square test. Multicollinearity between model parameters was evaluated using variance inflation index. The optimal probability threshold was determined as the value that maximised Youden’s index. The intercept for the logistic model was adjusted so that the probability threshold for a positive result was set to 50%. Non-parametric percentile bootstrap was performed with 2000 samples to assess the stability of the coefficients and calculate 95% confidence intervals for all performance metrics^12^. A two-sided p-value < 0.05 was considered statistically significant.

## Results

### Study population

The patient inclusion flowchart is shown in Figure 1. From 1,670 patients with chest pain who were managed through the RACC during the time period, 589 underwent SE plus had an available digital clinic ECG, and after exclusions, 292 patients were included as the final study population. The baseline characteristics are presented in Table 1. The mean age was 58±14 years and 150 (51%) were males. The majority of patients had a left ventricular ejection fraction (LVEF) > 50%, 6 (2%) had a mildly reduced LVEF (40–50%), and none had an LVEF <40%. The mean HEART score was 3±1, reflecting patients had a low-to-intermediate 6-week risk of MACE^10^. Both SE and ExECG were negative for inducible myocardial ischemia in 240 patients (82%), forming the negative SE group. Inducible myocardial ischemia was present on SE in 24 patients (8%), forming the positive SE group. The final subgroup were those with a negative SE but a positive ExECG positive (29 patients (10%), forming the discordant ExECG group. Patients in the positive SE and discordant ExECG groups were older than those in the negative group (p=0.01), otherwise the three groups were similar in baseline characteristics.

**Figure 1.**
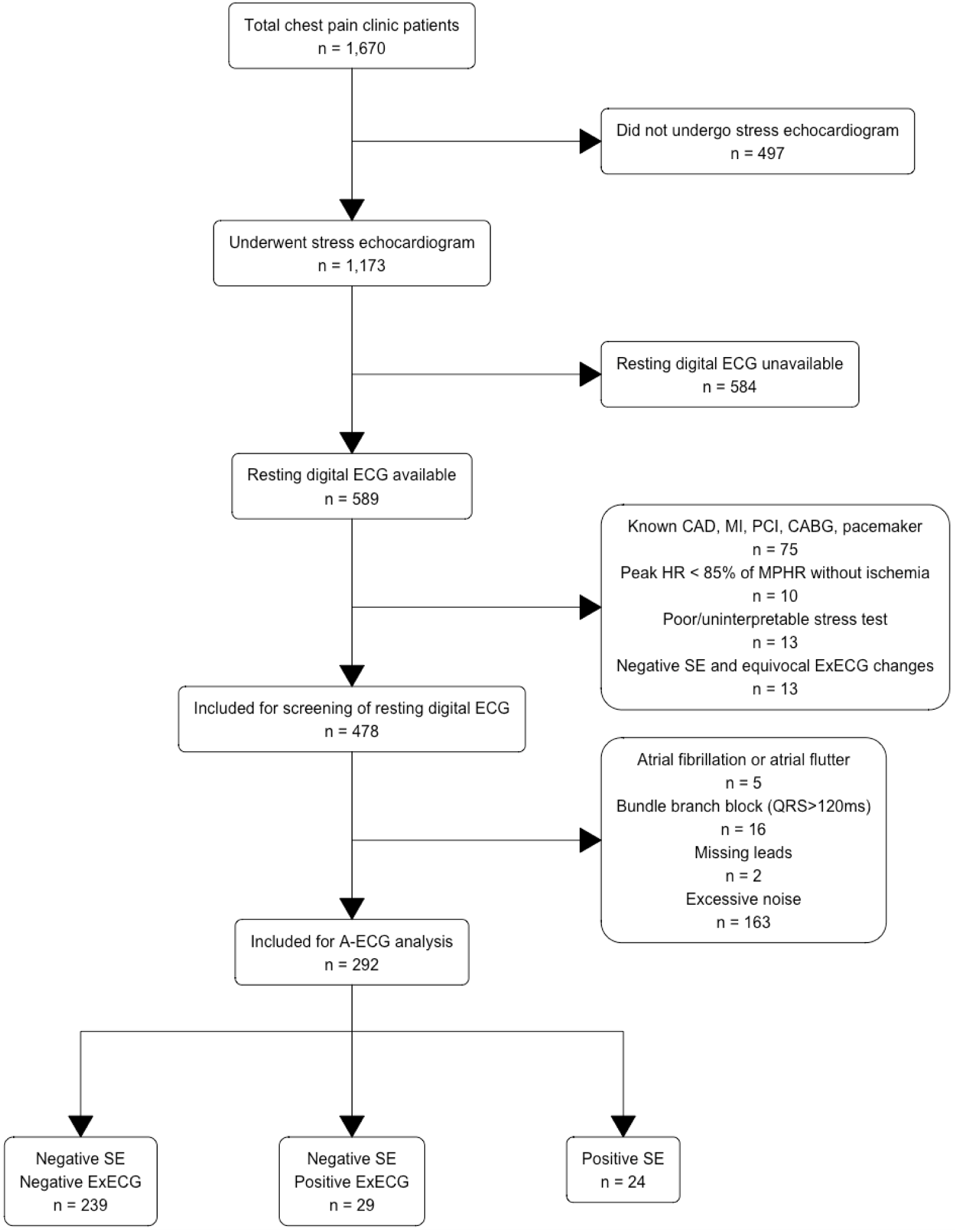
Patient inclusion flow chart and stress testing results. Abbreviations: ECG = electrocardiogram; SE = stress echocardiography; ExECG = exercise electrocardiography; A-ECG = advanced electrocardiography; CAD = coronary artery disease; MI = myocardial infarction; PCI = percutaneous coronary intervention; CABG = coronary artery bypass grafting; MPHR = maximum predicted heart rate (220-age); Equivocal ExECG changes = horizontal/down-sloping ST segment depression < 1 millimetre

**Table 1.**
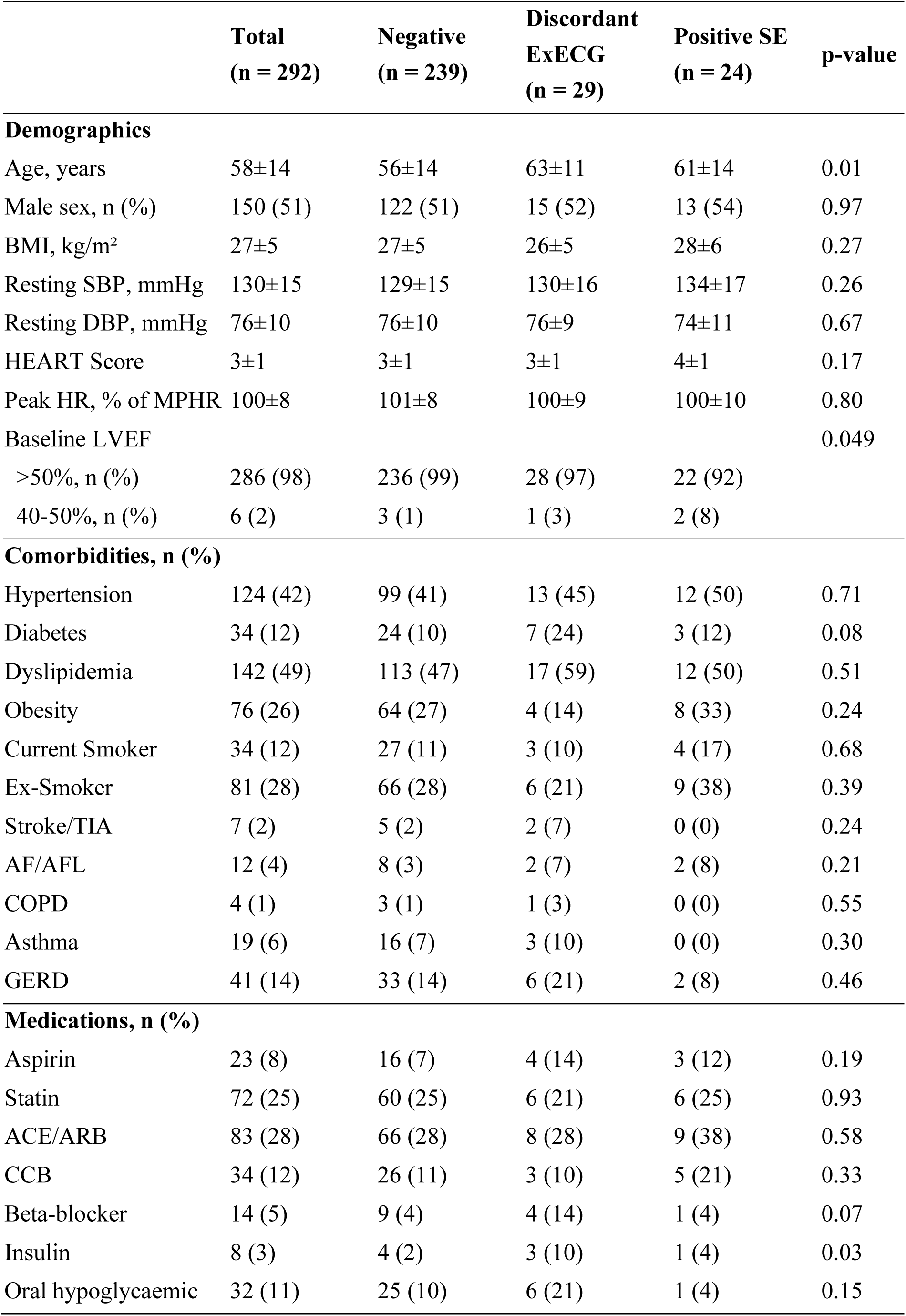
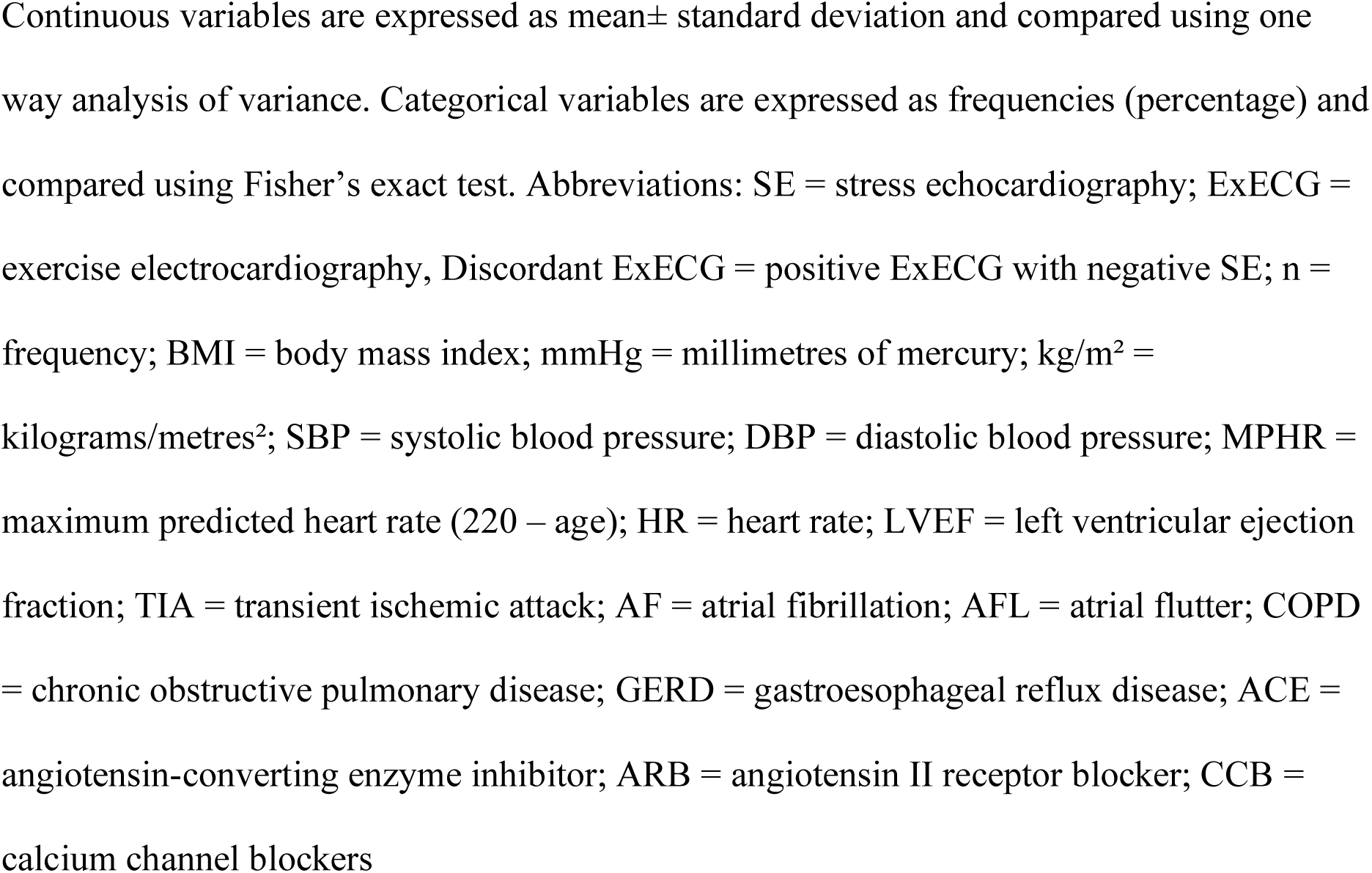
Patient demographics stratified by stress testing result.

### Development of the A-ECG Score

The best 3-parameter A-ECG score for predicting inducible myocardial ischemia by SE consisted of one conventional ECG measure and two advanced ECG measures, as follows: (1) the natural log of the R wave duration in lead V6 (R Dur V6), (2) the normalised 4th singular value of the T wave (4^th^ T Eigen), and (3) the azimuth direction at the final eighth of the spatial ST-T loop (ST-T Azimuth). All included measures were minimally correlated by variance inflation factor (< 1.1) and had stable coefficients across bootstrapping. These measures are compared between the negative and positive groups in Table 2. The regression coefficients are reported in *Table 3*. After re-arranging the logit function, the A-ECG score for inducible ischemia on SE was calculated as:

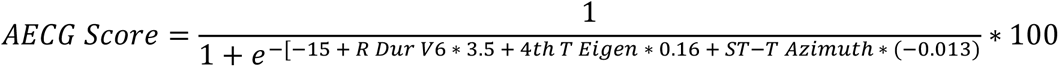

**Table 2.**
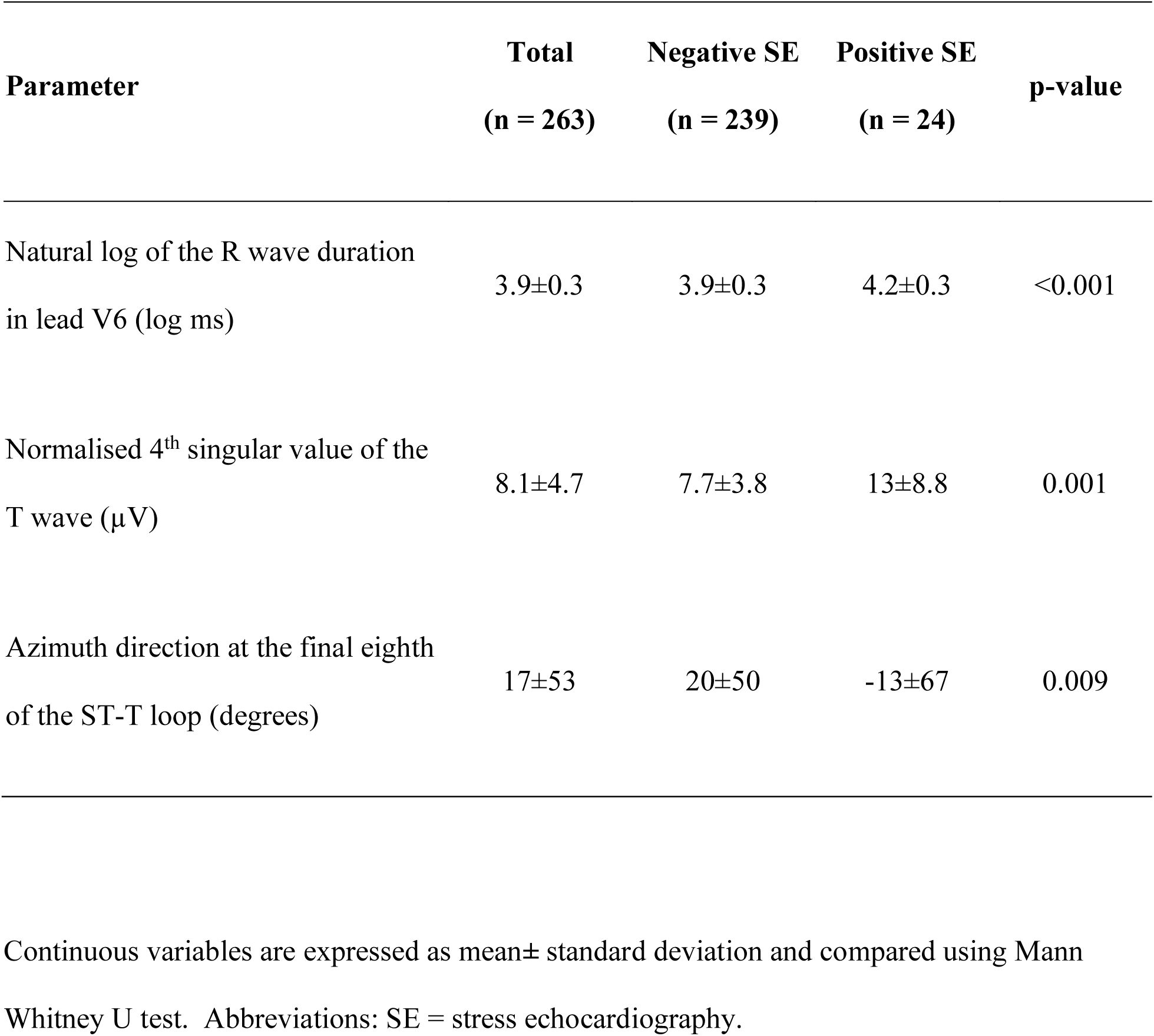
Parameter Means with Mann Whitney U test.

**Table 3.** Regression coefficients for the logistic model.

|  | **Coefficient** | **Chi-square** | **p-value** |
| --- | --- | --- | --- |
| **Intercept** | -15 |  | - |
| Natural log of the R wave duration in lead V6 (log ms) | 3.5 | 17 | <0.001 |
| Normalised 4<sup>th</sup> singular value of the T wave ($\mu$ V) | 0.16 | 17 | <0.001 |
| Azimuth direction at the final eighth of the ST-T loop (degrees) | -0.013 | 9.3 | 0.002 |

Coefficients are assessed using the likelihood-ratio chi-square test.

### A-ECG Score Performance

The diagnostic performance of the optimal 3-parameter score for detecting inducible myocardial ischemia on SE is presented in Table 4 and Figure 2. The probability cut-off with the highest Youden’s index was 0.06, and this was scaled to 50% by adjusting the intercept of the model. Of the 24 patients in the positive SE group, 2 (8%) had scores below the adjusted threshold of 50%, representing false negatives. 78 out of the 239 patients (33%) in the negative group had scores above 50%, representing false positives. As an example, Figure 3 compares the resting ECGs between two representative patients with high and low A-ECG scores for inducible myocardial ischemia on SE.

**Figure 2.**
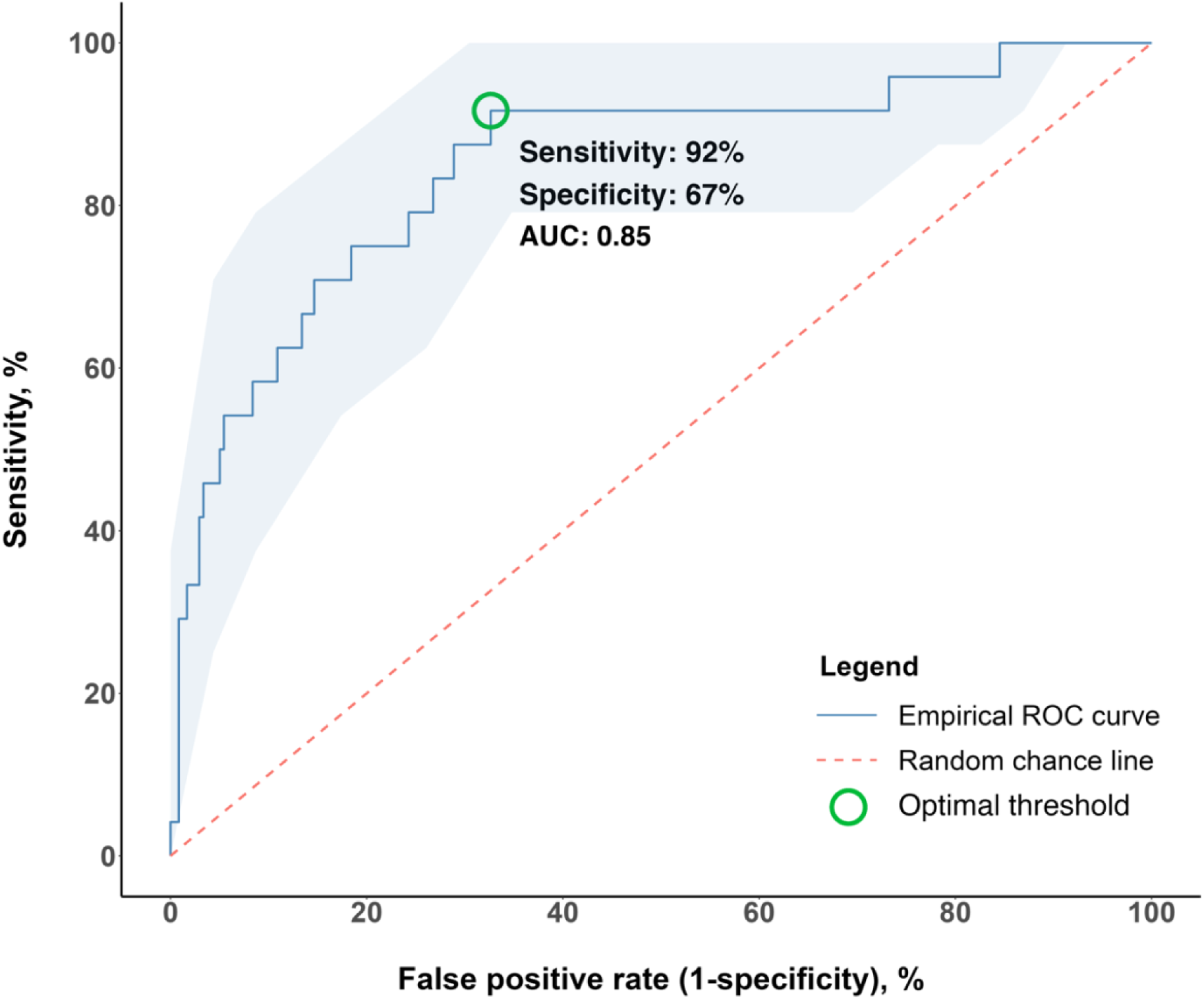
Empirical receiver-operating characteristic (ROC) curve and its 95% confidence interval for the 3-parameter score trained for inducible myocardial ischemia on stress echocardiography. The random chance line is given by: true positive rate (sensitivity) = false positive rate (1-specificity), representing an ROC curve with no discrimination. Abbreviations: ROC = receiver**-**operating characteristic

**Figure 3.**
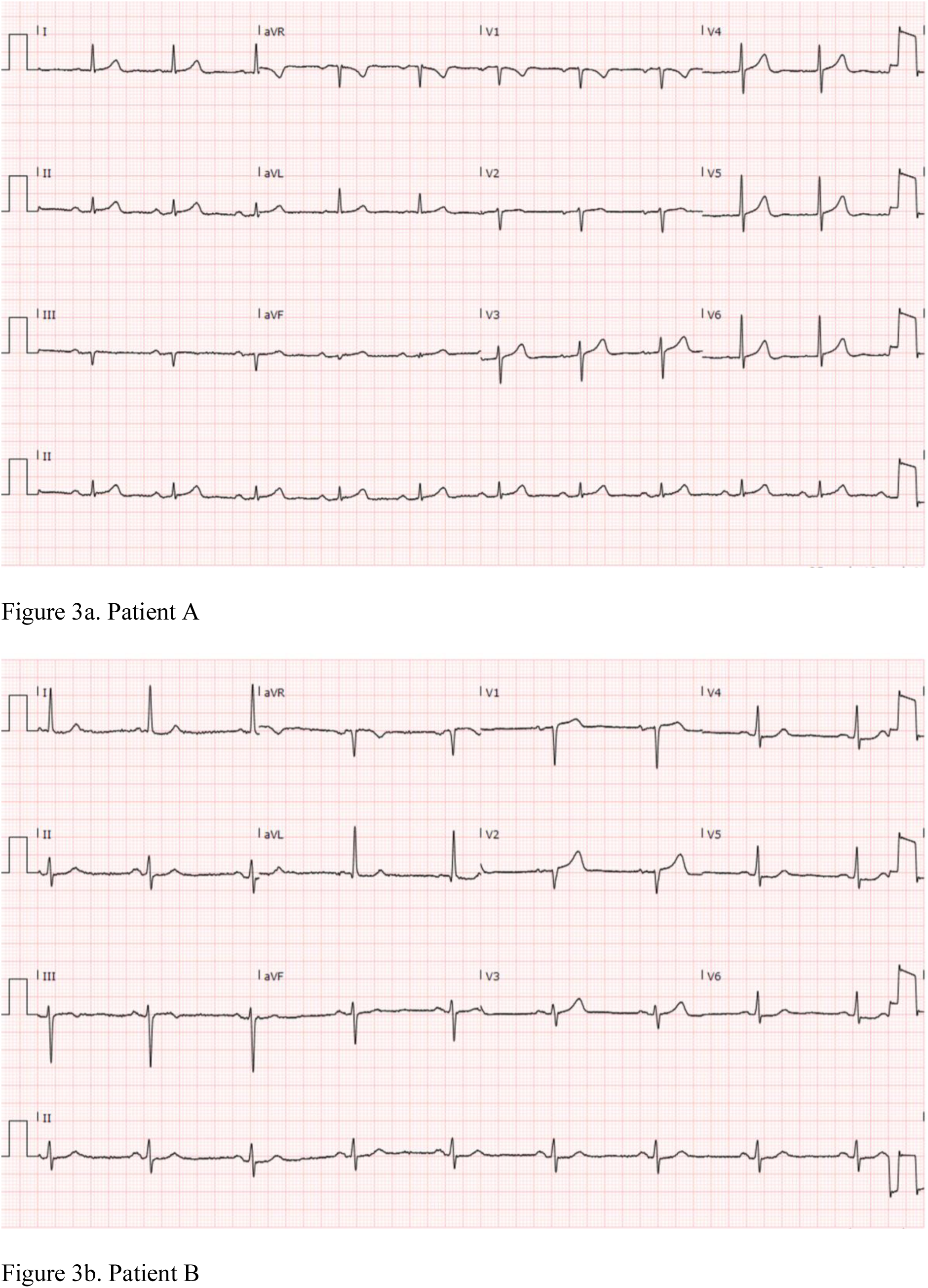
Comparison of A-ECG results of two representative patients. Both Patient A and Patient B had no signs of ischemia on resting ECGs and normal resting echocardiography. On stress echocardiography, Patient A showed no evidence of inducible myocardial ischemia, with a low corresponding A-ECG score for inducible myocardial ischemia (2%). Patient B developed hypokinesis of the basal-to-mid left ventricular inferior wall, with a high corresponding A-ECG score for inducible myocardial ischemia (95%).

**Table 4.**
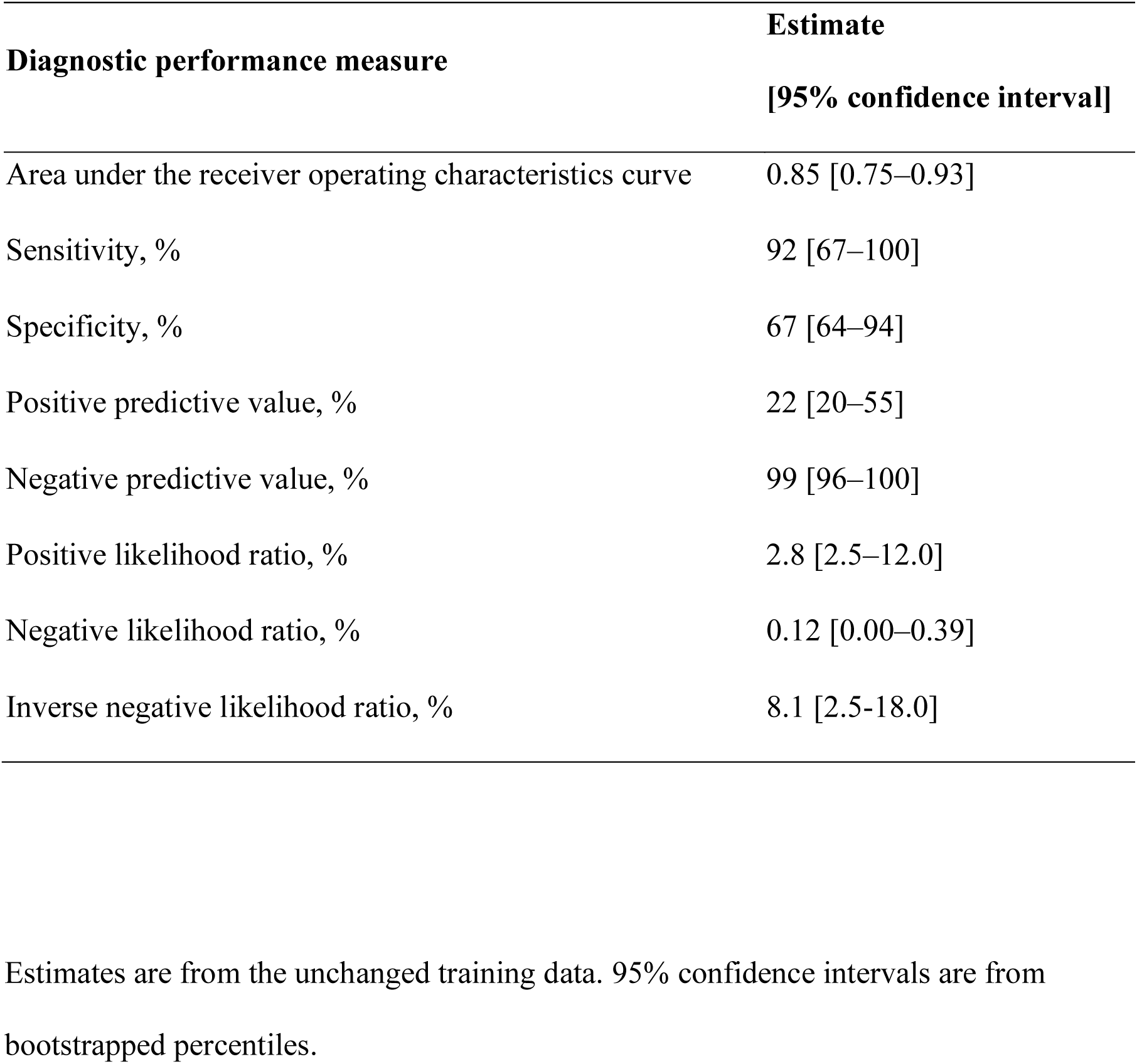
Diagnostic performance of 3-parameter score for inducible myocardial ischemia on stress echocardiography.

| Diagnostic performance measure | Estimate<br>[95% confidence interval] |
| --- | --- |
| Area under the receiver operating characteristics curve | 0.85 [0.75–0.93] |
| Sensitivity, % | 92 [67–100] |
| Specificity, % | 67 [64–94] |
| Positive predictive value, % | 22 [20–55] |
| Negative predictive value, % | 99 [96–100] |
| Positive likelihood ratio, % | 2.8 [2.5–12.0] |
| Negative likelihood ratio, % | 0.12 [0.00–0.39] |
| Inverse negative likelihood ratio, % | 8.1 [2.5-18.0] |

Estimates are from the unchanged training data. 95% confidence intervals are from bootstrapped percentiles.

**Table 5.** Clinical and ECG information comparison between Patient A and Patient B.

|  | Patient A | Patient B |
| --- | --- | --- |
| **Myocardial ischemia on SE** |  |  |
| True result | Negative | Positive |
| A-ECG score | 95% | 2% |
| **A-ECG measures** |  |  |
| Natural log of the R wave duration in lead V6 (log ms) | 3.6 | 3.8 |
| Normalised 4<sup>th</sup> singular value of the T wave (μV) | 5.5 | 17.7 |
| Azimuth direction at the final eighth of the ST-T loop (degrees) | 150.0 | -153.78 |
| **Clinical Information** |  |  |
| Age, years | 66-70 | 81-85 |
| Sex | Female | Female |
| BMI, kg/m<sup>2</sup> | 22.3 | 22.6 |
| Resting SBP, mmHg | 140 | 142 |
| Resting DBP, mmHg | 70 | 70 |
| HEART Score | 3 | 3 |
| Peak HR, % of MPHR | 106 | 85 |

Abbreviations: SE = stress echocardiography; A-ECG = advanced electrocardiography; BMI = body mass index; SBP = systolic blood pressure; DBP = diastolic blood pressure; HR = heart rate; MPHR = maximum predicted heart rate (220 – age)

## Discussion

This study demonstrates that a 3-parameter A-ECG score applied to a standard resting 12-lead ECG has a good AUC and excellent negative predictive value and inverse negative likelihood for ruling out inducible myocardial ischemia on SE. This is the first study to investigate the ability of A-ECG to detect inducible myocardial ischemia on SE, and thus could be used as a simple test to help triage low-to-intermediate risk patients into needing a SE or not.

SE has been validated as an excellent non-invasive test for the diagnosis of significant CAD and is widely performed to investigate stable chest pain^1,2^. Although invasive angiography is the gold standard for the diagnosis of CAD, SE provides independent prognostic information^13,14^. For these reasons, SE was chosen as the primary outcome to train the logistic model.

### Explainable A-ECG Measures

Measures of waveform complexity are obtained through singular value decomposition, a method of dimensionality reduction. In healthy patients, most of the variance can be explained from a single eigenvalue (the 1^st^ eigenvalue). In patients with cardiac disease, higher numbered eigenvectors account for greater voltage as the signal becomes more complex^15^. The normalised 4^th^ singular value of the T wave was included in our model, being elevated in patients with positive SE. This indicates elevated T wave complexity and abnormal repolarisation. Large studies have shown measures of waveform complexity to predict incident heart failure and cardiovascular mortality^8,16^.

The QRS complex reflects depolarisation of the ventricular myocardium. The natural log of the R wave duration in lead V6 was included in our model, being elevated in patients with positive SE. This may reflect conduction delay caused by ischemia to the conduction system and myocardium. In general populations, QRS duration is independently associated with CAD, myocardial infarction and cardiovascular mortality^17,18^. Also, in patients referred to stress echocardiography for suspected CAD, QRS duration is an independent predictor of cardiac death and non-fatal infarction^19^.

Vectorcardiography (VCG) is the representation of the heart’s electrical activity in three spatial planes^20^. In A-ECG analysis, the VCG is derived from the standard 12-lead ECG using validated methods^21^, providing ready access to hundreds of additional VCG parameters. Our model contained one VCG-derived measure: the azimuth direction at the final eighth of the spatial ST-T loop, which was more positively directed in patients with negative SE. The mean values for this measure in our negative SE cohort are similar to those previously reported for healthy men^22^, whereas those in our positive SE cohort likely diverge from normal^23^. However, the mechanisms behind these observed directional differences in T wave orientation require further elucidation.

### A-ECG Model Performance

The high NPV and low PPV observed in our model is inherent to the low prevalence of outcome events. If used in a similar population to ours, patients with chest pain that receive an A-ECG score < 50% are highly unlikely to exhibit inducible myocardial ischemia on SE and thus we suggest would not require a SE or further testing for CAD. The poorer PPV indicates that a positive A-ECG score does not guarantee inducible ischemia on SE. At face value, this would greatly reduce its clinical utility. However, the utility of the A-ECG score is not to replace SE, but to optimise its testing population. In our study cohort, if the 163 out of 263 patients with an A-ECG score < 50% were discharged without referral to SE, only 2 of these (1.2%) would have gone on to test positive. Doing so would greatly reduce the level of unnecessary testing and associated costs while missing very few patients that would otherwise test positive.

### Comparison to Clinical Risk Scores

Current triage of patients to SE depends on risk of major adverse cardiovascular events and pre-test probability for CAD based on clinical assessment. Clinical risk scores such as the Diamond-Forrester and Duke clinical cores were developed to estimate pre-test probability for CAD based on easily obtained clinical findings^24,25^. However, these scores have been shown to overestimate pre-test probability for CAD in contemporary populations^26,27^. This may contribute to the decrease in incidence of positive SE results over the last decades^3,4^.

There is a growing sentiment that the level of cardiac testing is inappropriately high, incurring unnecessary costs to patients and the healthcare system^28^. One study aimed to identify the clinical factors that specifically predicted inducible myocardial ischemia on nuclear stress testing^29^. Known CAD, typical angina and reduced LVEF were identified as the strongest independent predictors^29^. Our study offers an alternative method of estimating risk of CAD, by using advanced and conventional ECG parameters. We did not incorporate any clinical variables into our model in order to specifically assess the predictive power of A-ECG variables alone. While clinical predictors are robust due to their extensive real-world usage and validation^26^, A-ECG measures have a theoretical basis alongside proven clinical utility in many large studies^7,8,16,30^.

### Comparison to Conventional ECG

The resting conventional ECG is readily available and forms a component of many clinical risk scores for CAD. Previous studies have attempted to quantify the diagnostic potential of the resting conventional ECG for CAD outside of STEMI. One cross-sectional study demonstrated that patients with resting ECG abnormalities have a higher prevalence of subclinical atherosclerosis by coronary calcium score^31^. Another study determined that resting ECG abnormalities defined by the Minnesota code could predict myocardial ischemia on stress perfusion magnetic resonance imaging with a hazard ratio of 2.5 (95% CI: 1.4-4.4)^32^. Our study did not use pre-determined ECG abnormalities to predict myocardial ischemia. Rather, features selection methods were used to select the optimal parameters from a large pool of conventional and advanced ECG parameters. As a result, our A-ECG score can predict myocardial ischemia on SE even on resting ECGs that demonstrate no remarkable findings to the naked eye. An example of this is demonstrated in Figure 3.

### Comparison to Other A-ECG Scores

The excellent performance of A-ECG for predicting inducible myocardial ischemia on SE is in alignment with previous studies which have demonstrated its ability to distinguish various cardiac conditions^6,9,33–35^. A-ECG has been shown to outperform optimized pooled criteria from the conventional ECG. A 7-parameter A-ECG score has been shown to have a sensitivity of 89% and specificity of 94% for predicting pooled CAD, left ventricular hypertrophy, and left ventricular systolic dysfunction^6^. Also, within our same cohort of chest pain patients, A-ECG has been validated with a high specificity of 86% for ruling out significant stenosis on CCTA^9^. A recent study showed that A-ECG can accurately diagnose apical hypertrophic cardiomyopathy while also distinguishing from other cardiac disease using linear discriminant analysis^35^. This was able to accurately identify CAD with an AUC of 0.99 and specificity of 98%^35^.

### Comparison to Artificial Intelligence

There is an emerging body of research addressing the medical applications of machine learning, with the ECG being a prime target due to its raw digital data that is available in large numbers for training^36^. Multivariable logistic regression itself is a form of supervised machine learning used for regression and classification. Artificial intelligence (AI) usually refers to unsupervised deep learning, which is beneficial in that it does not require human selection of parameters that is limited by current scientific understanding^36^. Studies have shown that AI can accurately predict various cardiac conditions from a resting ECG^37,38^. However, the prevailing issue with unsupervised AI is that its decision process cannot be interpreted by humans – a so-called “black box”. Consequently, there exists a sentiment that AI may not be appropriate for guiding medical decisions^39^. By comparison, the A-ECG score from the current study is comprised of known A-ECG measures and can therefore be considered transparent or explainable, when compared to unsupervised AI methods.

### Clinical Implications

The current study demonstrates the utility of a resting 12-lead A-ECG score to screen low-to-intermediate risk chest pain patients for those that would exhibit inducible myocardial ischemia on SE. The conventional 12-lead ECG is routinely performed in the emergency department and outpatient clinics as a quick, cost-effective, and simple test for chest pain. An A-ECG score inherits these advantages plus provides diagnostic information that cannot be discerned by the human eye. Automatic A-ECG analysis of a resting 12-lead ECG can quickly identify the probability of testing positive for inducible myocardial ischemia on SE. This analysis can be applied to ECGs acquired from any vendor that stores a digital raw data file that is accessible to the user. Hence, the addition of an A-ECG score may significantly improve triage to SE and reduce unnecessary testing in this patient population.

### Limitations

The study was performed retrospectively from a single centre and may not be applicable to other patient populations. It also contains a limited sample size, particularly in the outcome group. There exists the possibility of overfitting, whereby the model trains on noise specific to our patient sample. Consequently, the diagnostic performance of the A-ECG score may decrease when applied to another sample. Due to the limited data, it was not feasible to split patients into a training and test group. Instead, the model was trained on all available data and resampled using non-parametric bootstrapping to obtain 95% confidence intervals. Also, the number of parameters incorporated into the model was limited to 3 to prevent overfitting. We did not correlate the SE results with any further tests such as invasive or CT coronary angiography.

Future studies should aim to test the reproducibility of the A-ECG score in an independent validation cohort, the clinical implementation in an emergency department setting, and assessing short and long term clinical and economical outcomes.

## Conclusions

A 3-parameter resting A-ECG score has a good AUC and excellent inverse negative likelihood ratio for detecting inducible myocardial ischemia on SE. This supports the use of A-ECG assessment on a resting 12-lead ECG to effectively identify patients with chest pain at low-to-intermediate risk that do or do not require further investigation for CAD. However, further larger prospective validation in independent cohorts is warranted.

## Data Availability

All data produced in the present study are available upon reasonable request to the authors

## Authorship

**Kevin X Yang:** Software, Validation, Formal Analysis, Investigation, Data curation, Writing - Original Draft, Writing – Review & Editing, Visualisation. **Johan von Scheele:** Software, Validation, Formal analysis, Investigation, Data curation, Writing – Original Draft, Writing – Review & Editing, Visualisation. **Maren Maanja:** Software, Formal analysis, Writing – Review & Editing. **Daniel E Loewenstein:** Software, Formal analysis, Resources, Writing – Review & Editing. **Todd T Schlegel:** Software, Formal analysis, Resources, Writing – Review & Editing. **Martin Ugander:** Conceptualisation, Methodology, Resources, Writing - Review & Editing, Supervision, Project Administration, **Rebecca Kozor:** Conceptualisation, Methodology, Resources, Data Curation, Writing - Review & Editing, Supervision, Project Administration.

